# Early detection of prostate gland and breast cancer risk based on routine check-up data using survival analysis trees for left-truncated and right-censored data

**DOI:** 10.1101/2021.01.11.21249491

**Authors:** Dan Coster, Eyal Fisher, Shani Shenhar-Tsarfaty, Tehillah Menes, Shlomo Berliner, Ori Rogowski, David Zeltser, Itzhak Shapira, Eran Halperin, Saharon Rosset, Malka Gorfine, Ron Shamir

## Abstract

**Objective:** To predict breast cancer (BC) and prostate gland cancer (PGC) risk among healthy individuals by analyzing routine laboratory measurements, vital signs and age.

**Materials and Methods:** We analyzed electronic medical records of 20,317 healthy individuals who underwent routine checkups, encompassing more than 600 parameters per visit, and identified those who later developed cancer. We developed a novel ensemble method for risk prediction of multivariate time series data using a random forest model of survival trees for left truncated and right-censored data.

**Results:** Using cross-validation, our method predicted future PGC and BC 6 months before diagnosis, achieving an area under the ROC curve of 0.62±0.05 and 0.6±0.03 respectively, better than standard random forest, Cox-regression model and a single survival tree. Our method can complement existing screening tests such as clinical breast examination and mammography for BC, and help in detection of subjects that were missed by these tests.

**Discussion:** Computational analysis of results of routine checkups of healthy individuals can improve the detection of those at risk of cancer development.

**Conclusion:** Our method may assist in early detection of breast and prostate gland cancer.

## BACKGROUND AND SIGNIFICANCE

Early detection of cancer is crucial for providing appropriate care to the patient and can improve both prognosis and survival [1–5]. The current detection strategies use specific screening tests that require substantial resources, e.g., serum Prostate-Specific Antigen (PSA) level for Prostate Gland Cancer (PGC), mammography, an X-ray modality, for detecting early signs of Breast Cancer (BC), and clinical breast examination (CBE), a physical examination to recognize abnormalities in the breast [6]. Other approaches to assess cancer risk use models, e.g. Gail’s model [7,8], BRCAPRO [9], IBIS [10] and BOADICEA [11] for BC risk, and the Prostate Cancer Prevention Trial Risk Calculator (PCPTRC) for PGS risk [12]. Both models use a few clinical parameters and are not based on routine laboratory measurements, and their performance is relatively limited [13].

Machine learning algorithms can improve screening models in two major directions. One approach is utilizing advanced algorithms to improve the performance of the existing tests, for example, deep learning models for analyzing mammography [14–16], machine learning models for optimizing Gail’s model parameters [16], and improving the PGC risk score based on longitudinal PCPTRC results [17]. Another approach aims to develop new cancer risk prediction tools based on historical medical records of patients, collected as part of routine care in Electronic Medical Records (EMR). Such models were suggested for lung cancer [18], colorectal cancer [19], and Acute Myeloid Leukemia [20], among others. Moreover, advanced genetic methods are also employed for screening, mostly using polygenic risk scores [21]. Our objective is to develop new models for both BC and PGC based on EMR data collected from healthy individuals in routine periodic checkups, using techniques from machine learning and survival trees.

Survival trees were first introduced by Gordon and Olsen [22]. The basic concept is to create a decision tree where each node contains a survival curve of the corresponding subgroup of individuals. The node splitting criterion usually aims to maximize the difference in survival between the subgroups of the daughter nodes or the within-node homogeneity. Most survival tree methods addressed right-censored data and time-independent covariates. Incorporating time-varying covariates in survival trees was first introduced in [23] by introducing the ‘pseudo-object’ concept, which we describe later.

Several later methods of constructing survival trees for time-dependent covariates used this concept [24–27] or others [28–30]. Another common approach for analyzing time-dependent covariates is the Cox-regression model [31,32].

Several ensemble methods for survival trees analysis were suggested for time-independent covariates [33,34]. Random survival forests (RSF), introduced by Ishwaran [35] and combined the concepts of Breiman’s random forest [36,37], survival trees and the log-rank test as the splitting criterion. An extension of RSF is the utilization of conditional inference trees, which use hypothesis testing to select the splitting covariates and also as a stopping criterion [38], among other improvements that were examined [39,40].

We considered the problem of predicting survival probability over time. Our objective was to create a model based on subjects’ time-dependent covariates obtained in routine laboratory tests and to predict the fully personalized survival function for each subject based on the last available measurement values. We developed a novel method called TVsuRF (Time-Varying SUrvival Random Forest) for this goal. TVsuRF is the first ensemble method based of survival trees for time-dependent covariates that implements the ‘pseudo-object’ concept. Moreover, our method is the first to use the conditional inference trees in that setting.

Today, screening tests in the healthy population are used to identify individuals with cancer without symptoms, but these tests are costly, labor-intensive, and suffer from low accuracy. Our method aims to utilize existing clinical measurements of healthy individuals to predict the risk of BC and PGC, the most common cancers among females and males, respectively. To the best of our knowledge, this is the first risk score that is based on routine laboratory measurements proposed for these cancer types.

## MATERIALS AND METHODS

### Dataset

We analyzed data from routine checkups of individuals at the Tel-Aviv Medical Center Inflammation Survey (TAMCIS), Tel-Aviv Sourasky Medical Center, Israel. Participants were men and non-pregnant women with no active current malignant or infectious disease who chose to be tested and signed an informed consent form. In each visit, the subject underwent a comprehensive medical history evaluation, a complete physical examination, blood and urine tests, vital signs measurements, an electrocardiogram, an exercise stress test, and a respiratory function test. Data were summarized in structured EMR. Some individuals had multiple visits during several years. We conducted a retrospective analysis of the TAMICS EMR data collected between November 2001 and February 2017. Our study covered 20,271 adults (age ≥ 18). The study was reviewed and approved by the Institutional Review Board (Approval no. 02-049-Tlv).

### Cancer Registry

TAMICS participants who later developed cancer were identified (using their national IDs) in the Israeli National Cancer Registry (INCR), which records all cancer cases in Israel. INCR contains for each case the cancer type (ICD9 code) and diagnosis date, and we used all cancer diagnoses until January 1^st^ 2016. **Supplementary Figure 1** shows the number of patients in the cohort with each cancer type. We focused on the two cancer types with the largest number of cases: BC for females and PGC for males. Patients who had a different type of cancer prior to diagnosis of BC or PGC were excluded.

### Exclusion & Inclusion Criteria

#### Inclusion criteria

All individuals surveyed in TAMICS who had birth and visit dates documented were included (number of individuals *n*_*p*_= 20,271, number of visits *n*_*v*_= 50,497). Of those, individuals with cancer diagnosis according to INCR were identified (*n*_*p*_= 1,547, *n*_*v*_=3,999), along with their cancer type (see **Figure 1**).

**Figure 1:**
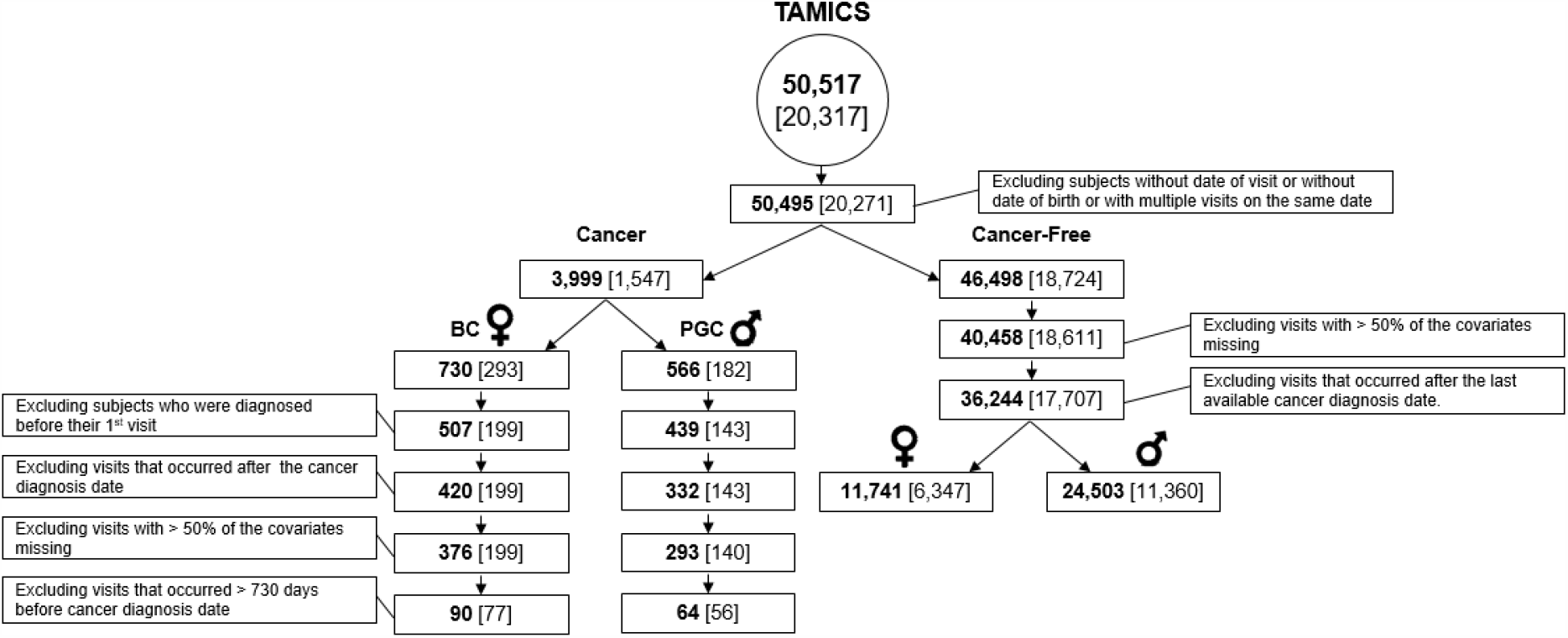
Study design. The bold number is the number of TAMICS visits; the number of individuals appears in parentheses.

#### Cases

Females whose cancer type was BC (*n*_*p*_= 293, *n*_*v*_=730) or males whose cancer type was PGC (*n*_*p*_= 182, *n*_*v*_=566).

#### Controls

Individuals who did not have any cancer diagnosis (*n*_*p*_= 18,724, *n*_*v*_= 46,498).

#### Exclusion criteria

Our analysis was based on data from single visits, so exclusion was done per individual and visit.

#### Cases

Individuals whose cancer diagnosis date was before their first TAMICS visit (BC: *n*_*p*_= 94, *n*_*v*_=223, PGC: *n*_*p*_= 39, *n*_*v*_=127). Visits that occurred after the cancer diagnosis date (BC: *n*_*v*_=87, PGC: *n*_*v*_=107). Visits where more than 50% of the covariates were missing (BC: *n*_*v*_=44, PGC: *n*_*v*_=39). Visits that occurred > 730 days before the cancer diagnosis date (BC: *n*_*p*_= 122, *n*_*v*_=286, PGC: *n*_*p*_= 84, *n*_*v*_=229).

#### Controls

Visits where more than 50% of the covariates were missing (*n*_*p*_= 113 individuals and *n*_*v*_=6,040 visits excluded). Visits that occurred after the last day of reports in INCR (*n*_*p*_= 934, *n*_*v*_=4,214). We split the cancer-free group into male (*n*_*p*_= 11,360, *n*_*v*_=24,503), and female (*n*_*p*_= 6,347, *n*_*v*_=11,741) subgroups.

### Data Extraction and Feature Choices

We used only features that were available for more than 80% of the individuals. The missing values were imputed using Predictive-Mean-Matching on age [41] using the *mice* package [42].

For BC risk prediction we used 20 covariates (**Table 1**) that include demographic parameters such as age and BMI, along with Complete Blood Count (CBC), since BC is a systemic disease that affects the immune system and its progression is expected to be reflected in the CBC results. For PGC risk prediction, we added 28 covariates that include the Basic Metabolic Panel (BMP), Lipids, Vital Signs, and more. (**Table 2**)

**Table 1.** Characteristics of the BC, BC-free and Matched BC-free groups. Values are mean ± SD. MW: p-value of the Mann–Whitney test, T-test: p-value of the Student t-test. All p-values were Bonferroni corrected for multiple hypotheses. Baso – basophils; EOS – eosinophils; Hmt – hematocrit, Hgb-hemoglobin; Lym – lymphocytes; MCH-mean corpuscular hemoglobin; MCHC-mean corpuscular hemoglobin concentration; MCV - mean corpuscular volume; Mono-monocytes; MPV-mean platelet volume; Neu – neutrophils; RBC – red blood cells; PLT – platelets; RDW - red cell distribution width; WBC – white blood Cells; BMI - body mass index

|  | **BC** |  |  | **BC-Free** |  |  | **Matched BC-Free** |  |  | **BC vs. BC-Free**<br>**P-value** |  | **BC vs. Matched**<br>**BC-Free P-value** |  |
| --- | --- | --- | --- | --- | --- | --- | --- | --- | --- | --- | --- | --- | --- |
| **Parameter** | **Visits** | **Subjects** | **Mean±STD** | **Visits** | **Subjects** | **Mean±STD** | **Visits** | **Subjects** | **Mean±STD** | **T-test** | **MW** | **T-test** | **MW** |
| **Baso (%)** | 90 | 77 | 0.63±0.33 | 11,739 | 6,347 | 0.58±0.29 | 5,883 | 3,635 | 0.59±0.3 | 1 | 1 | 1 | 1 |
| **Eos (%)** | 90 | 77 | 2.61±1.73 | 11,738 | 6,347 | 2.5±1.84 | 5,882 | 3,635 | 2.54±1.78 | 1 | 1 | 1 | 1 |
| **Hmt (%)** | 90 | 77 | 39.06±2.62 | 11,741 | 6,347 | 38.59±2.81 | 5,884 | 3,635 | 38.88±2.86 | 1 | 1 | 1 | 1 |
| **Hgb (g/dL)** | 90 | 77 | 13.2±0.96 | 11,740 | 6,347 | 13.15±0.96 | 5,883 | 3,635 | 13.24±0.96 | 1 | 1 | 1 | 1 |
| **Lym (%)** | 90 | 77 | 30.71±8.26 | 11,739 | 6,347 | 30.75±7.17 | 5,883 | 3,635 | 30.99±7.2 | 1 | 1 | 1 | 1 |
| **Lym (K/μL)** | 90 | 77 | 2.13±0.76 | 11,734 | 6,347 | 2.04±0.57 | 5,880 | 3,635 | 2.01±0.56 | 1 | 1 | 1 | 1 |
| **MCH (pg)** | 90 | 77 | 29.8±2.27 | 11,740 | 6,347 | 29.95±2.04 | 5,884 | 3,635 | 30.04±2.06 | 1 | 1 | 1 | 1 |
| **MCHC(g/dL)** | 90 | 77 | 33.85±0.86 | 11,740 | 6,347 | 34.11±0.98 | 5,884 | 3,635 | 34.08±1.05 | 0.114 | **0.049** | 0.344 | 0.159 |
| **MCV (fl)** | 90 | 77 | 87.99±5.62 | 11,741 | 6,347 | 87.75±5.06 | 5,884 | 3,635 | 88.1±5.09 | 1 | 1 | 1 | 1 |
| **Mono (%)** | 90 | 77 | 6.88±1.45 | 11,739 | 6,347 | 6.97±1.91 | 5,883 | 3,635 | 7.12±1.71 | 1 | 1 | 1 | 1 |
| **Mono (K/μL)** | 90 | 77 | 0.48±0.16 | 11,734 | 6,347 | 0.46±0.15 | 5,880 | 3,635 | 0.46±0.13 | 1 | 1 | 1 | 1 |
| **MPV (fl)** | 87 | 74 | 9.19±0.97 | 11,312 | 6,234 | 9.01±1.07 | 5,688 | 3,559 | 9.01±1.08 | 1 | 1 | 1 | 1 |
| **Neu (K/μL)** | 90 | 77 | 4.23±1.42 | 11,734 | 6,347 | 4.06±1.37 | 5,880 | 3,635 | 3.95±1.33 | 1 | 1 | 1 | 0.739 |
| **RBC (M/μL)** | 90 | 77 | 4.45±0.35 | 11,740 | 6,347 | 4.4±0.34 | 5,883 | 3,635 | 4.42±0.35 | 1 | 1 | 1 | 1 |
| **Neu (%)** | 90 | 77 | 59.16±8.63 | 11,739 | 6,347 | 59.21±8.17 | 5,883 | 3,635 | 58.75±8.16 | 1 | 1 | 1 | 1 |
| **PLT (K/μL)** | 90 | 77 | 262.67±52.95 | 11,740 | 6,347 | 263.17±61.56 | 5,884 | 3,635 | 261.35±61.31 | 1 | 1 | 1 | 1 |
| **RDW (%)** | 90 | 77 | 13.42±1.26 | 11,741 | 6,347 | 13.25±1.06 | 5,884 | 3,635 | 13.29±1.02 | 1 | 1 | 1 | 1 |
| **WBC (K/μL)** | 90 | 77 | 7.07±1.84 | 11,741 | 6,347 | 6.77±1.7 | 5,884 | 3,635 | 6.63±1.66 | 1 | 1 | 0.538 | 0.379 |
| **BMI (kg/m<sup>2</sup>)** | 83 | 71 | 25.9±4.74 | 11,273 | 6,057 | 25.45±4.72 | 5,574 | 3,445 | 26.23±4.63 | 1 | 1 | 1 | 1 |
| **Age (Years)** | 90 | 77 | 53.46±7.97 | 11,741 | 6,347 | 47.16±10.56 | 5,884 | 3,635 | 53.2±7.66 | **< 0.0001** | **< 0.0001** | 1 | 1 |

**Table 2.** Characteristics of the PGC, PGC-free and Matched PGC-free groups. Values are mean ± SD. MW: p-value of the Mann–Whitney test, T-test: p-value of the Student t-test. P-values are Bonferroni corrected for multiple hypotheses. DBP – diastolic blood pressure; SBP – systolic blood pressure; Temp – body temperature; BUN - blood urea nitrogen ; GGT - gamma-glutamyl transferase; ALP - alkaline phosphatase; LDH – lactate dehydrogenase; Urine SG-urine specific gravity; Urine PH – PH stick for urine test.

|  | <u>PGC</u> |  |  | <u>PGC-Free</u> |  |  | <u>Matched PGC-Free</u> |  |  | <u>PGC vs. PGC-Free</u><br><u>P-value</u> |  | <u>PGC vs. Matched</u><br><u>PGC-Free P-value</u> |  |
| --- | --- | --- | --- | --- | --- | --- | --- | --- | --- | --- | --- | --- | --- |
| Parameter | Visits | Subjects | Mean±STD | Visits | Subjects | Mean±STD | Visits | Subjects | Mean±STD | T-test | MW | T-test | MW |
| Baso (%) | 64 | 56 | 0.57±0.26 | 24,382 | 11,344 | 0.54±0.27 | 6,080 | 3,320 | 0.54±0.27 | 1 | 1 | 1 | 1 |
| Eos (%) | 64 | 56 | 2.51±1.32 | 24,382 | 11,344 | 2.86±1.87 | 6,080 | 3,320 | 2.92±1.86 | 1 | 1 | 0.809 | 1 |
| Hmt (%) | 64 | 56 | 43.65±2.8 | 24,390 | 11,344 | 43.73±2.7 | 6,083 | 3,320 | 43.71±2.87 | 1 | 1 | 1 | 1 |
| Hgb (g/dL) | 64 | 56 | 14.93±0.97 | 24,390 | 11,344 | 14.94±0.94 | 6,083 | 3,320 | 14.9±1 | 1 | 1 | 1 | 1 |
| Lym (%) | 64 | 56 | 27.52±6.89 | 24,382 | 11,344 | 29.79±6.74 | 6,080 | 3,320 | 28.58±6.78 | 0.537 | 1 | 1 | 1 |
| Lym (K/μL) | 63 | 55 | 1.8±0.53 | 24,369 | 11,269 | 1.98±0.56 | 6,079 | 3,290 | 1.93±0.59 | 0.597 | 0.748 | 1 | 1 |
| MCH (pg) | 64 | 56 | 30.33±1.67 | 24,389 | 11,344 | 30.17±1.66 | 6,083 | 3,320 | 30.46±1.76 | 1 | 1 | 1 | 1 |
| MCHC (g/dL) | 64 | 56 | 34.23±0.79 | 24,389 | 11,344 | 34.21±0.89 | 6,083 | 3,320 | 34.13±0.92 | 1 | 1 | 1 | 1 |
| MCV (fl) | 64 | 56 | 88.57±4.14 | 24,390 | 11,344 | 88.18±4.28 | 6,083 | 3,320 | 89.25±4.46 | 1 | 1 | 1 | 1 |
| Mono (%) | 64 | 56 | 8.06±1.97 | 24,382 | 11,344 | 7.99±1.8 | 6,080 | 3,320 | 8.21±1.86 | 1 | 1 | 1 | 1 |
| Mono (K/μL) | 63 | 55 | 0.54±0.17 | 24,370 | 11,269 | 0.53±0.16 | 6,079 | 3,290 | 0.56±0.16 | 1 | 1 | 1 | 1 |
| MPV (fl) | 63 | 55 | 8.87±1.22 | 23,498 | 11,257 | 8.85±1.02 | 5,899 | 3,289 | 8.84±1.05 | 1 | 1 | 1 | 1 |
| Neu (K/μL) | 63 | 55 | 4.18±1.37 | 24,368 | 11,269 | 4.01±1.28 | 6,078 | 3,290 | 4.14±1.29 | 1 | 1 | 1 | 1 |
| RBC (M/μL) | 64 | 56 | 4.93±0.38 | 24,387 | 11,344 | 4.97±0.36 | 6,083 | 3,320 | 4.9±0.38 | 1 | 1 | 1 | 1 |
| Neu (%) | 64 | 56 | 61.34±8.05 | 24,382 | 11,344 | 58.82±7.52 | 6,080 | 3,320 | 59.75±7.52 | 0.767 | 1 | 1 | 1 |
| PLT (K/μL) | 64 | 56 | 244.08±80.53 | 24,389 | 11,344 | 238.68±55.85 | 6,083 | 3,320 | 233.5±55.56 | 1 | 1 | 1 | 1 |
| RDW (%) | 64 | 56 | 13.34±0.86 | 24,389 | 11,344 | 13.01±0.79 | 6,083 | 3,320 | 13.2±0.84 | 0.190 | 0.138 | 1 | 1 |
| WBC (K/μL) | 64 | 56 | 6.71±1.66 | 24,390 | 11,344 | 6.75±1.64 | 6,083 | 3,320 | 6.87±1.67 | 1 | 1 | 1 | 1 |
| Pulse (bpm) | 59 | 53 | 69.95±14.05 | 23,053 | 10,896 | 68.68±11.86 | 5,591 | 3,155 | 68.14±11.7 | 1 | 1 | 1 | 1 |
| DBP (mmHg) | 59 | 53 | 81.05±8.26 | 23,331 | 10,896 | 78.66±8.63 | 5,672 | 3,155 | 80.71±8.55 | 1 | 1 | 1 | 1 |
| SBP (mmHg) | 59 | 53 | 131.44±15.59 | 23,326 | 10,896 | 125.1±14.32 | 5,671 | 3,155 | 131.08±15.48 | 0.142 | 0.099 | 1 | 1 |
| Spirometry (Score) | 56 | 50 | 0.34±0.48 | 22,563 | 10,716 | 0.39±0.49 | 5,435 | 3,080 | 0.4±0.49 | 1 | 1 | 1 | 1 |
| Temp. (C°) | 59 | 53 | 36.34±0.33 | 22,104 | 10,947 | 36.35±0.34 | 5,397 | 3,184 | 36.33±0.33 | 1 | 1 | 1 | 1 |
| BUN (mg/dL) | 61 | 55 | 16.34±3.75 | 24,056 | 11,003 | 15.36±3.67 | 6,027 | 3,195 | 16.37±4.15 | 1 | 1 | 1 | 1 |
| Chloride (mmol/L) | 60 | 54 | 104.05±2.53 | 24,015 | 10,920 | 103.52±2.42 | 6,023 | 3,160 | 103.64±2.56 | 1 | 1 | 1 | 1 |
| Creatinine(mg/dL) | 60 | 54 | 1.15±0.12 | 24,019 | 10,920 | 1.14±0.15 | 6,026 | 3,160 | 1.16±0.16 | 1 | 1 | 1 | 1 |
| GGT (U/L) | 60 | 54 | 27.57±23.54 | 23,993 | 10,920 | 25.07±22.42 | 6,018 | 3,160 | 26.36±22.21 | 1 | 1 | 1 | 1 |
| Glucose (mg/dL) | 61 | 55 | 100.18±21.96 | 24,059 | 11,003 | 92.58±16.83 | 6,030 | 3,195 | 97.51±19.7 | 0.457 | 0.002 | 1 | 1 |
| Potassium(mmol/L) | 60 | 54 | 4.45±0.35 | 24,019 | 10,920 | 4.35±0.37 | 6,025 | 3,160 | 4.37±0.38 | 1 | 0.511 | 1 | 1 |
| Albumin (g/L) | 60 | 54 | 44.8±2.13 | 24,014 | 10,920 | 45.52±2.32 | 6,022 | 3,160 | 44.82±2.27 | 0.599 | 1 | 1 | 1 |
| Globulin (g/L) | 60 | 54 | 27.12±3.67 | 23,995 | 10,920 | 28.12±3.2 | 6,017 | 3,160 | 27.98±3.25 | 1 | 1 | 1 | 1 |
| Phosphorus(mg/dL) | 60 | 54 | 3.16±0.39 | 24,012 | 10,920 | 3.23±0.44 | 6,022 | 3,160 | 3.16±0.43 | 1 | 1 | 1 | 1 |
| Calcium(mg/dL) | 60 | 54 | 9.35±0.43 | 24,011 | 10,920 | 9.32±0.42 | 6,021 | 3,160 | 9.27±0.43 | 1 | 1 | 1 | 1 |
| Uric Acid (mg/dL) | 60 | 54 | 6.19±1.12 | 23,995 | 10,920 | 6.09±1.1 | 6,016 | 3,160 | 6.17±1.14 | 1 | 1 | 1 | 1 |
| Sodium (mmol/L) | 60 | 54 | 141.82±2.91 | 24,019 | 10,920 | 141.19±2.53 | 6,025 | 3,160 | 141.09±2.58 | 1 | 1 | 1 | 1 |
| Protein (g/L) | 60 | 54 | 71.92±4.18 | 24,005 | 10,920 | 73.64±3.91 | 6,020 | 3,160 | 72.8±3.89 | 0.118 | 0.049 | 1 | 1 |
| Bilirubin (μmol/L) | 60 | 54 | 0.81±0.37 | 24,014 | 10,920 | 0.83±0.37 | 6,023 | 3,160 | 0.81±0.33 | 1 | 1 | 1 | 1 |
| ALP (U/L) | 59 | 53 | 63.85±17.3 | 23,214 | 10,840 | 64.64±17.54 | 5,850 | 3,131 | 64.48±17.57 | 1 | 1 | 1 | 1 |
| LDH (U/L) | 60 | 54 | 323.6±44.04 | 24,013 | 10,920 | 317.76±55.91 | 6,022 | 3,160 | 324.77±55.11 | 1 | 1 | 1 | 1 |
| Triglycerides(mg/dL) | 63 | 56 | 126.63±56.12 | 24,207 | 11,260 | 123.48±73.12 | 6,044 | 3,289 | 127.33±70.01 | 1 | 1 | 1 | 1 |
| HDL (mg/dL) | 63 | 56 | 47.42±11.16 | 24,182 | 11,260 | 49.81±10.67 | 6,036 | 3,289 | 50.63±11.54 | 1 | 1 | 1 | 0.810 |
| LDL (mg/dL) | 63 | 56 | 114.54±28.54 | 24,095 | 11,260 | 115.78±29.83 | 6,023 | 3,289 | 113.03±30.3 | 1 | 1 | 1 | 1 |
| Cholesterol (mg/dL) | 63 | 56 | 188.27±35.1 | 24,204 | 11,260 | 190.14±34.74 | 6,043 | 3,289 | 189.01±35.08 | 1 | 1 | 1 | 1 |
| Troponin (ng/dL) | 63 | 56 | 4.11±1.04 | 24,141 | 11,260 | 3.94±0.97 | 6,026 | 3,289 | 3.86±0.9 | 1 | 1 | 1 | 1 |
| Urine PH | 64 | 56 | 6.14±0.89 | 24,134 | 11,344 | 6.13±0.82 | 6,014 | 3,320 | 6.1±0.81 | 1 | 1 | 1 | 1 |
| Urine SG | 64 | 56 | 1.01±0.01 | 24,112 | 11,344 | 1.01±0.05 | 6,005 | 3,320 | 1.01±0.05 | 1 | 1 | 1 | 1 |
| BMI (kg/m<sup>2</sup>) | 62 | 54 | 27.34±3.29 | 23,543 | 11,177 | 26.88±3.74 | 5,729 | 3,266 | 27.74±3.65 | 1 | 1 | 1 | 1 |
| Age (Years) | 64 | 56 | 59.61±6.33 | 24,471 | 11,344 | 47.13±10.78 | 6,102 | 3,320 | 59.24±5.77 | < 0.0001 | < 0.0001 | 1 | 1 |

## Methods

### Preliminaries

Consider a dataset of *N* subjects, where for each of them data from one or more visits were recorded. Subject *i* had *M*^*i*^ visits at times 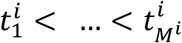. The *d* covariates measured at time 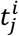 are denoted by the vector 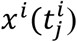 (For simplicity, we assume that all covariates were recorded in every visit). Note that covariates can be either time-dependent or time-independent (static). Hence, 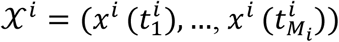 summarizes the longitudinal data of subject *i* The last time point subject *i* was at risk, which can be either failure or censoring time, is 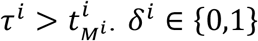 denotes if the subject experienced a censoring (*δ*^*i*^ = 0) or failure event (*δ*^*i*^ = 1) at time *τ*^*i*^. Hence, the full data can be summarized by the set of triplets 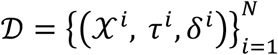 (**Supplementary Figure 2A**). *x*^*i*^(*t*) denotes the data of subject *i* that were measured until time *t*, i.e., 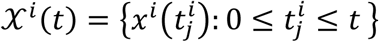. We assume time homogeneity so that w.l.o.g. we can shift times per subject to set 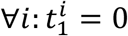, i.e., all first visits were at time 0 (**Supplementary Figure 2B**). We also assume that the age of the subject at each visit is one of the covariates.

Our model aims to estimate the probability for being free of the failure event (the cancer diagnosis) at least until time *t* based on the patient’s covariates at the latest visit before that time. That is, let 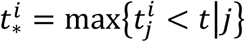. We wish to estimate the survival function:

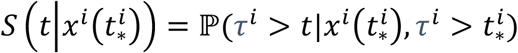

In order to model the time-dependent covariates, we transform the data following [23]. We split the data of each subject into disjoint intervals 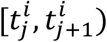 and we assume that the covariates *x*^*i*^(*t*_*j*_) are constant in the interval (**Supplementary Figure 2C)**. In that manner, we consider *t*_*j*_ as the left-truncation time. If 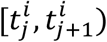 is not the last interval of subject *i* then we view time 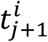 as censoring time. We denote the pseudo-object of the *j*^*th*^ interval of subject *i* as 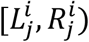 where:

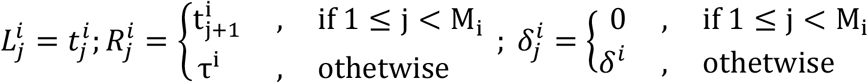

Hence, the transformation is:

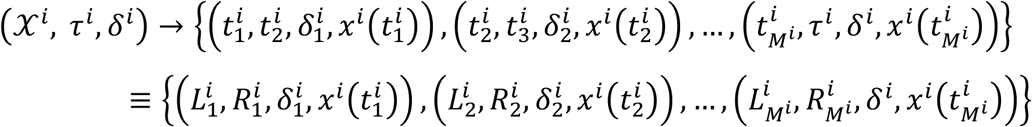

Each pseudo-interval is therefore possibly left-truncated and/or censored. The standard Kaplan-Meier (KM) estimator of the survival function can now be generalized for left truncation right-censored (LTRC) data [43], as follows. Assume that there were *D* failure events and they occurred at distinct times *t*_1_ < ⋯ < *t*_*D*_. We denote by *Y*_*j*_ the number of pseudo-objects at risk at time 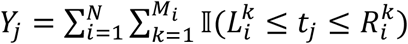 i.e., the number of individuals who entered the study before time *t*_*j*_ and did not experience a failure or censoring event until *t*_*j*_. *d*_*j*_ is defined as the number of patients that experienced a failure event at time *t*_*j*_ and due to our prior assumption *d*_*j*_ = 1. The KM estimator is defined as a step function with jumps at observed failure times:

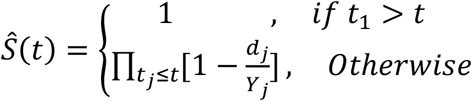

The survival probability will be calculated in a step ahead prediction manner - we calculate the probability of a patient in time *t* to experience failure in the next time window Δ*t* given its covariates at time *t*, namely ℙ (*τ*^*i*^ < *t* + Δ*t, δ*_*i*_ = 1|*τ*^*i*^ > *t, x*_*i*_(*t*)).

### Survival tree construction

We now describe the construction of the survival tree for pseudo-objects data. For simplicity, we will just call them objects. (**Figure 2A**). Suppose we have the set of samples along with their covariates as described above, and we wish to use the survival information to build a decision tree. We use the framework of conditional inference trees [38], a class of decision trees that employs a statistical hypothesis test based on permutations in order to select optimal variables and their thresholds. This process is different from common decision tree construction (see **Supplementary Material 3)**, which usually selects the variable that maximizes an information measure (e.g. Gini or entropy).

**Figure 2:**
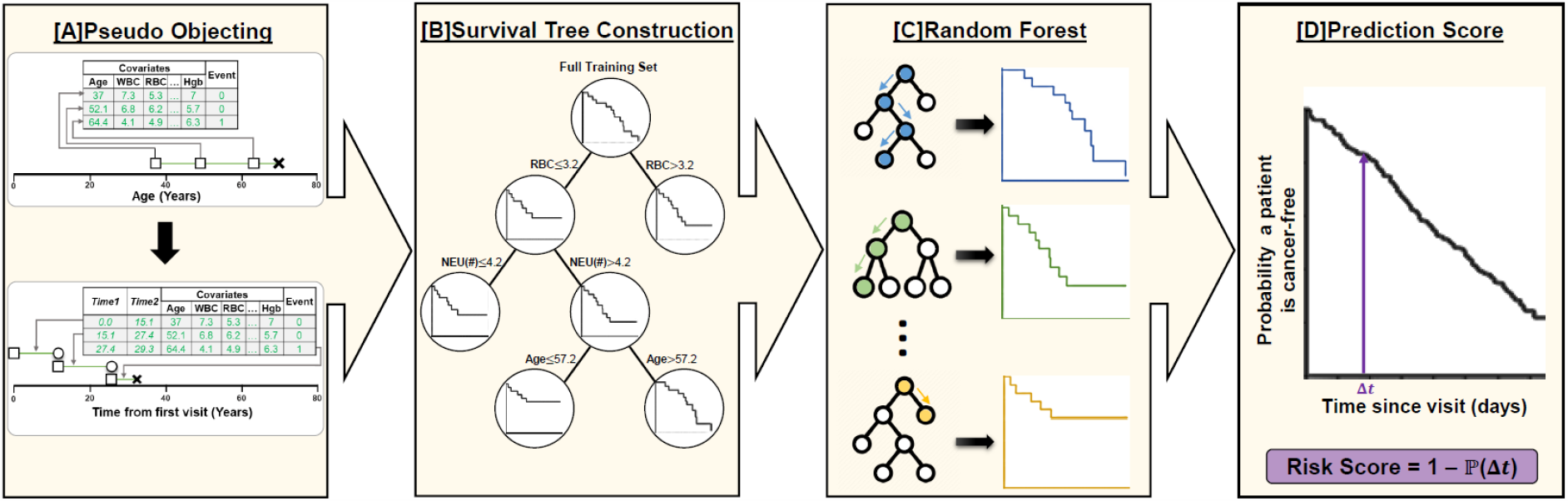
Model construction and evaluation. An illustration of the different parts of our model construction. [A] For each subject we transformed its data into pseudo objects and change the time axis to time from first visit. [B] An illustration of single survival tree construction [C] Generating 500 survival trees. [D] The trees are combined into a single unified model. Risk score calculation per each sample is based on averaged survival curve.

A covariate and a threshold value at a node split the node’s samples into two subsets, and each subset induces a survival curve. To compare the survival curves of the two subsets we use Pan’s permutations based hypothesis test [44], as suggested also in [27]. In every node, we test all possible covariates and thresholds, and the one that produces the split with the lowest p-value is selected. Notice that pseudo-objects created from the same subject can end in distinct sub-nodes.

The hypothesis test is based on creating an influence function that maps an object’s quadruplet (*L*_*i*_, *R*_*i*_, *δ*_*i*_, *x*_*i*_) into a scalar *U*_*i*_ which represents the contribution of sample *i* to the test statistic. We assume that (*l*_*i*_, *r*_*i*_) is the interval in which the true event lies, and denote its contribution to the statistic:

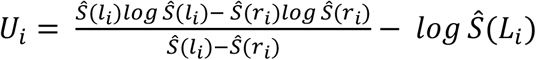

One can show that for failure event at time *t* (*δ*_*i*_ = 1)

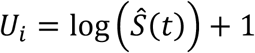

and for a right-censored observation at time *t* (*δ*_*i*_ = 0), assuming *Ŝ*(∞) = 0

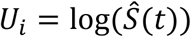

Now let *U*_1_, …, *U*_*N*_ be the scores of the samples corresponding to the parent node, and suppose *n* samples reside in the left child and *N* − *n* in the right. Write *X* = ∑_*left*_ *U*_*j*_. There are 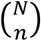 ways of choosing *n* out of the *N* scores and if *k* of these have a sum ≤ *X*, then assuming all partitions are equi-probable, the probability of obtaining a score of *X* is 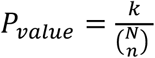. We estimate it using 1000 permutations.

The survival function *Ŝ*_*l*_(*t*) for node *l* is the Kaplan-Meier curve for the samples corresponding to that node. Let *C*_*l*_ be the set on indices of samples in node *l*, then:

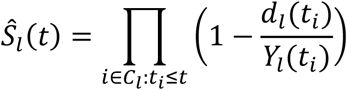

where *d*_*l*_(*t*_*i*_) is the number of failure events that occurred at time *t*_*i*_ in node *l* and *Y*_*l*_(*t*_*i*_) is the total number of objects at risk just before *t*_*i*_ in node *l*. (**Figure 2B, Figure 3**)

**Figure 3:**
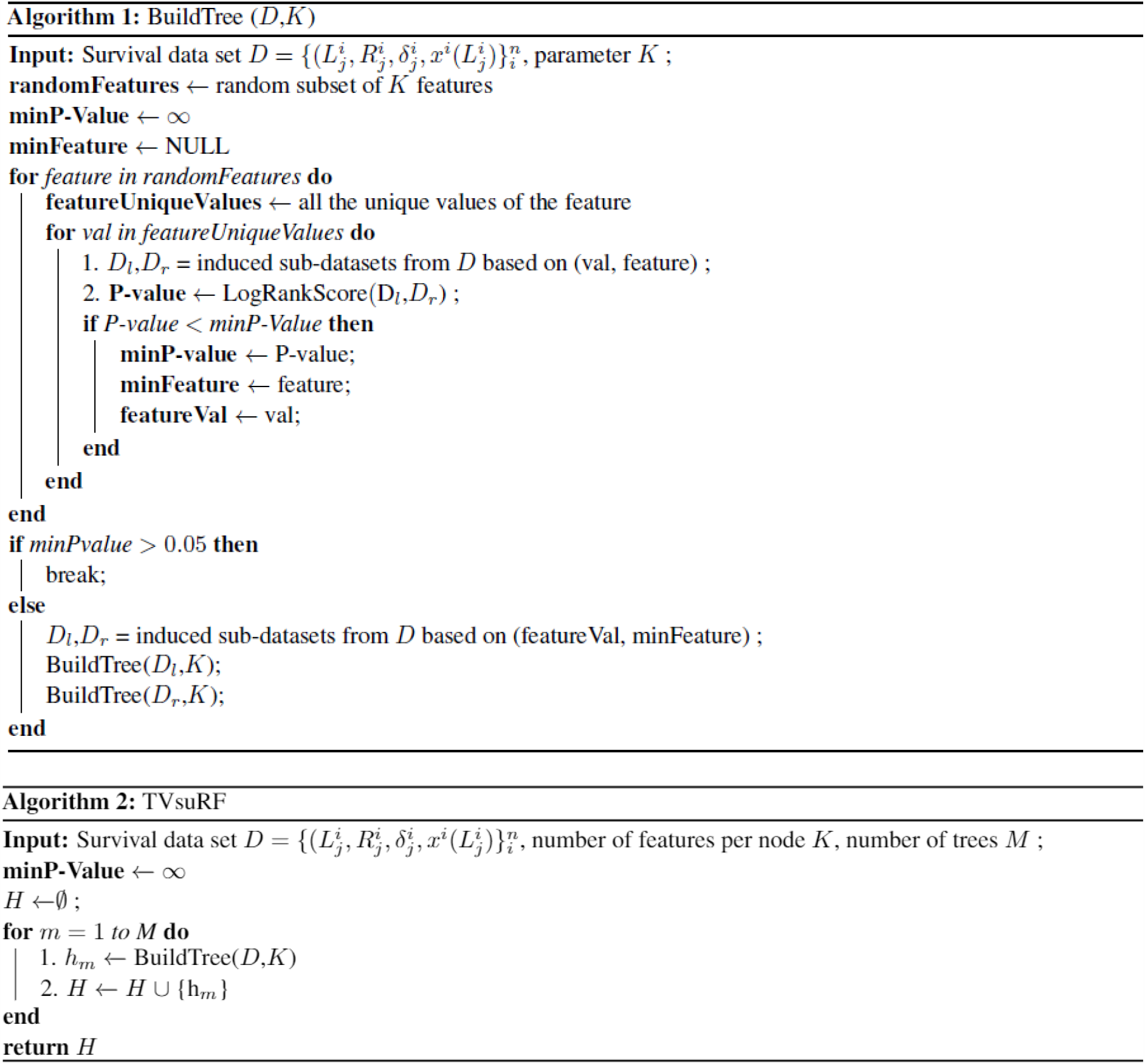
Algorithm 1: BuildTree Algorithm; Algorithm 2: TVsuRF Algorithm.

### Ensemble model

We create *M* = 500 survival trees. In each tree, at each internal node, we select at random 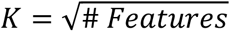 of the features and split the node according to the feature and threshold giving the least p-value for difference in survival, if that difference is significant (**Figure 3**). The predicted survival curve for a new subject *ω* is based on the data in all the leaves that *ω* ended in all the trees. Let 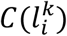 represent the set of indices of the subjects that are in the *i*^*th*^ leaf of the *k*^*th*^ tree and let 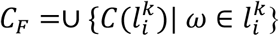 be the multiset of all the subjects in these leaves. If *d*_*i*_(*t*_*i*_) is the number of failure events in *C*_*F*_ at time *t*_*i*_ and *Y*_*i*_(*t*_*i*_) is the number of objects in *C*_*F*_ in risk at time *t*_*i*_, then the survival function of *ω* is (**Figure 2C**):

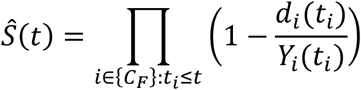

Our model constructs a Kaplan-Meier curve per each subject, producing a continuous risk score (RS) over time.

### Variable importance

We assessed the importance of each covariate in our model in two ways. In the first, we counted the fraction of internal nodes in all the trees that were associated with the covariate (i.e. the covariate was used to split these nodes). We call this fraction Vprop; higher Vprop indicates more importance. In the second approach, for each object, we replaced the values of the covariate by random values sampled independently from its original distribution, while keeping the other covariates in their true values, and recomputed the performance with the new data. The difference in the AUROC between the original and the modified data was computed and averaged over ten random assignments per each covariate on every fold of the 4-fold cross-validation [35]. We repeated this process 20 times and defined VIMP as the mean difference obtained. Again, higher VIMP indicates more importance.

### Comparison to BC screening tests

For a subset of the TAMICS females, we had data concerning BC screening. Mammography was available for 6,526 woman and Clinical Breast Exam (CBE) was available for 17,958. We excluded women with mutated BRCA genes, those who refused to conduct a CBE, lacked ID, had more than one record per visit or were diagnosed with another type of cancer (see **Supplementary Figure 4** for study design).

The result of the mammography was provided in free text written by the physician and transformed by us into binary labels (normal / abnormal) by natural language processing of the physician’s notes (see **Supplementary Material 5** for details). The CBE result was available as free text written by a physician and four binary values that represent an abnormal finding in the left/right breast or axilla. We considered the CBE result abnormal if one of the binary values was positive. In case that no values were reported, a breast cancer surgeon reviewed the physician’s text and decided if there was a positive finding.

We compared the recommendations that were done by these screening tests for BC to our predictions, in order to evaluate the added value of our approach. We binned the risk scores into deciles and the average risk score was calculated for each subject.

### Evaluation Approach

We used TVsuRF and several other models to predict BC and PGC risk on our cohorts. If a subject’s covariates were measured at time *t*, we aimed to predict cancer at time *t* + Δ*t*, for values of Δ*t* ranging between 183 and 730 days. Since there might be a delay between the cancer diagnosis time and the time it was reported to the cancer registry, we added *ϵ* =31 days to Δ*t*. The risk for patient *i* is thus:

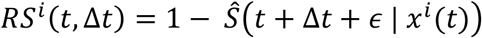

To evaluate the performance of this score for classification, we calculated the area under the receiver operator characteristic curve (AUROC), where the positive class is the set of individuals that were diagnosed with cancer during the next Δ*t* + *ϵ* days as suggested in

[45] (but excluding patients censored in [*t, t* + Δ*t* + *ϵ*]). We also estimated the area under the precision-recall (AUPR) curve.

We performed 20 iterations of 4-fold cross-validation, where in each iteration the partition of patients into folds was done at random. For each of the above measures, we calculated the average and standard deviation.

We compared our method to three others: (1) Cox regression model adapted to time-varying covariates [31,32], (2) single LTRC survival tree as in [27](denoted LTRCIT), and (3) RF model [36]. Since RF is a classification model, training for prediction was done separately for each time interval Δ*t*, and the class of a subject was positive if the diagnosis of cancer occurred during the next Δ*t* + *ϵ* days, and negative otherwise. We used 500 trees, and the ‘Gini’ index as a splitting rule, with the rest of the parameters at the default values in the *ranger* package [46] (**Figure 2D**).

In addition, we compared our method to a random survival forest (RSF) model that predicts a survival curve per sample. Since RSF was originally designed for handling time-independent covariates, we adapted it to our setting.

## RESULTS

### Breast Cancer

#### Dataset

Our cohort contained data on 6,424 women with a total of 11,831 visits to TAMICS. Out of those, 77 were diagnosed with breast cancer and had one or more visits less than 730 days before the diagnosis date (90 visits in total). These constituted the positive (BC) group. The covariates that were included in the model were CBC (18 parameters), age and BMI. The statistics of these values are summarized in **Table 1**.

Women in the positive group were significantly older on average than in the BC-free group and had significantly lower levels of mean corpuscular hemoglobin concertation (MCHC). To reduce the effect of age on our model, we created an age-matched cohort (‘Matched BC-Free’) using the approach of [47] (3,635 subjects, 5,884 visits). When comparing the BC and the Matched BC-free group (**Table 1**) none of the parameters was significantly different between the groups.

#### Prediction accuracy

The performance of each of the methods tested, for different time ranges, is summarized in **Figures 4A and 4B**. We also marked the AUROC of Gail’s breast cancer risk estimation for 5 years horizon as reported in [13]. TVsuRF had the highest AUPR on every time interval, and the highest AUROC on all intervals except one (though differences were not statistically significant) for 730 days, where Gail’s score was best. We also tested two versions of RSF and our model was better for time windows until 273 days in terms of AUPR and AUROC. (**Supplementary Figure 6**).

**Figure 4:**
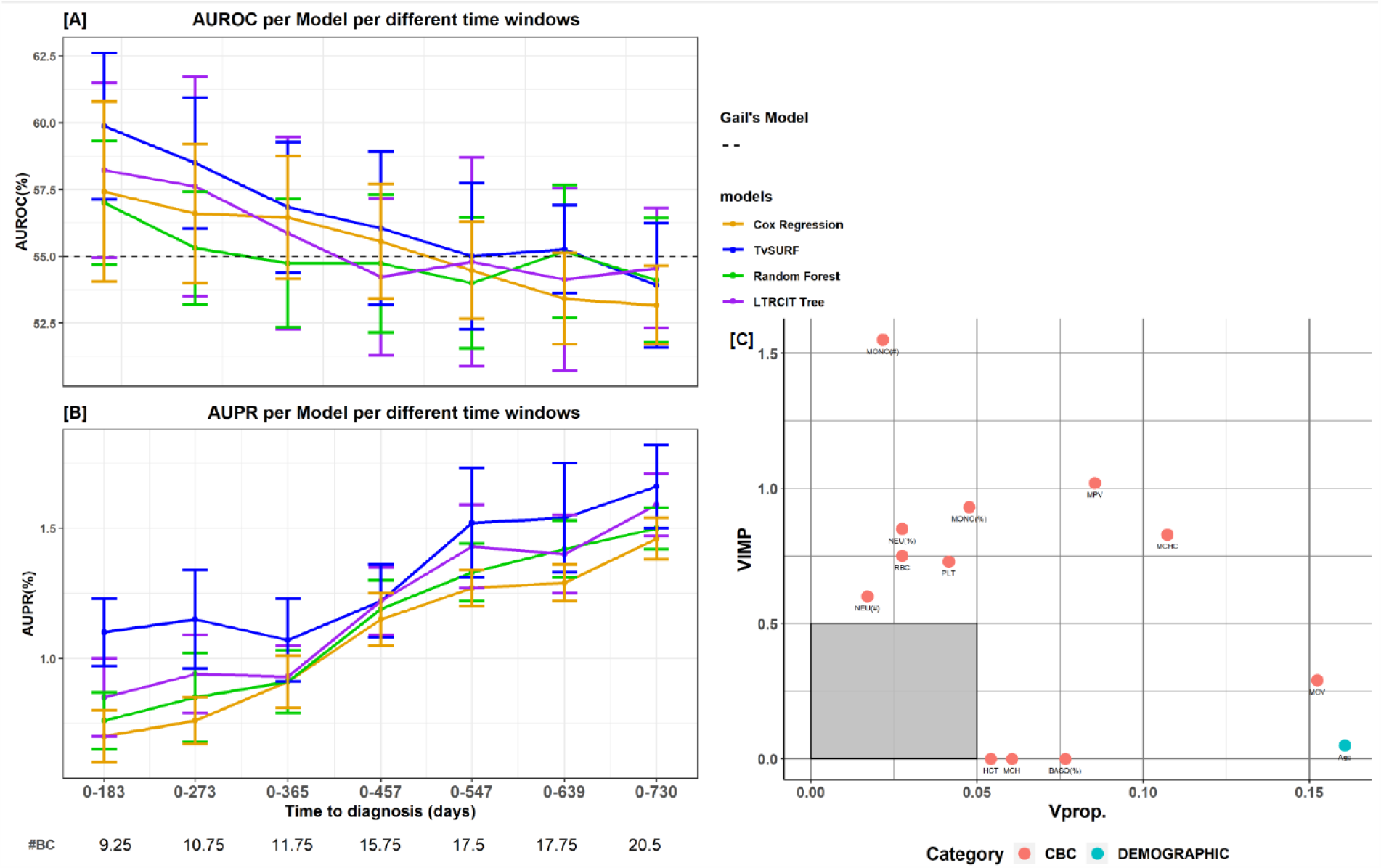
BC risk prediction and variable importance. [A] Performance (AUROC mean±SD) of five prediction models for different time intervals. The grey dashed line represents the (time-independent) AUROC reported for Gail’s Risk factor model [13]. [B] AUPR. The numbers below the x-axis labels are the average number of BC patients that were available across the cross-validation folds for each time interval. [C] Variable importance for model prediction in a 183-day window. Points indicate the different variables. The y-axis presents VIMP, the decrease in AUROC following random assignment of values to the variable. The x-axis plots Vprop, the variable’s inclusion frequency in the trees of the model. For both measures higher values indicate more importance. The color of a point represents the category of the parameter. Features of low importance (Vprop <0.05 and VIMP<0.5) are not shown.

#### Variable importance

**Figure 4C** summarizes the importance of variables in TVsuRF BC risk prediction model for a time window of 183 days. The variables mean corpuscular volume (MCV), monocytes (MONO), mean platelet volume (MPV), mean corpuscular hemoglobin concentration (MCHC) and age were most important in the TVsuRF model. The importance of immune system-related covariates such as MONO might correlate to the fact BC is an inflammatory and systemic disease.

#### Comparison to mammography and CBE

For every woman who underwent mammography or CBE in her checkup visits, we compared the results of the 730-day predictor, computed using data only from her latest visit. CBE had 29.1% sensitivity and 93.7% specificity, while TVsuRF had 12.5% sensitivity for the same specificity. Mammography sensitivity and specificity were 58.3% and 66.1%, and TVsuRF had 41.7% sensitivity for similar specificity. (Note that the results are not directly comparable, as mammography and CBE identify current malignancy and TVsuRF computes future disease risk.) The results in **Supplementary Figure 7** show the three predictions for women that were subsequently diagnosed with BC. Remarkably, the three women with the highest risk score estimated by our model were not detected by CBE, and one of them tested negative in mammography as well. In contrast, some of the women had lower risk scores but were detected by other screening tests.

### Prostate Gland Cancer

#### Dataset

This cohort consisted of 11,416 males who made a total of 24,567 visits to TAMICS. Out of them 56 were subsequently diagnosed with PGC and had 64 visits less than 730 days before the PGC diagnosis. We call this group the PGC subset. The covariates included in the model were CBC (20 parameters), basic metabolic panel data (BMP, 16 parameters), lipids (4 parameters), vital signs (5 parameters), urine tests (2 parameters), troponin, age and BMI. The characteristics of the covariates are summarized in **Table 2**. Since PGC individuals were significantly older than the PGC-free individuals, to reduce the effect of age on our model, we created an age-matched cohort (‘Matched PGC-Free’) of 3,320 subjects (6,083 visits) using the approach of [47] (**Table 2**). None of the covariates showed significant difference between the PGC and the Matched PGC-Free groups.

#### Prediction accuracy

**Figures 5A and 5B** show the results of five prediction methods, using the same comparison metrics as in the BC section. Our model had the highest AUROC in prediction window of 0-183 days and similar performance for intermediate size time windows. For windows of 547 days and longer, RF had the highest AUROC. In terms of AUPR, our model performed best in until 547 days and the advantage was significant in the windows of up to 273 days. When testing variants of RSF, TVsuRF had better performance on the prediction windows of 0-183 days, but less for longer time windows. (**Supplementary Figure 8**).

**Figure 5:**
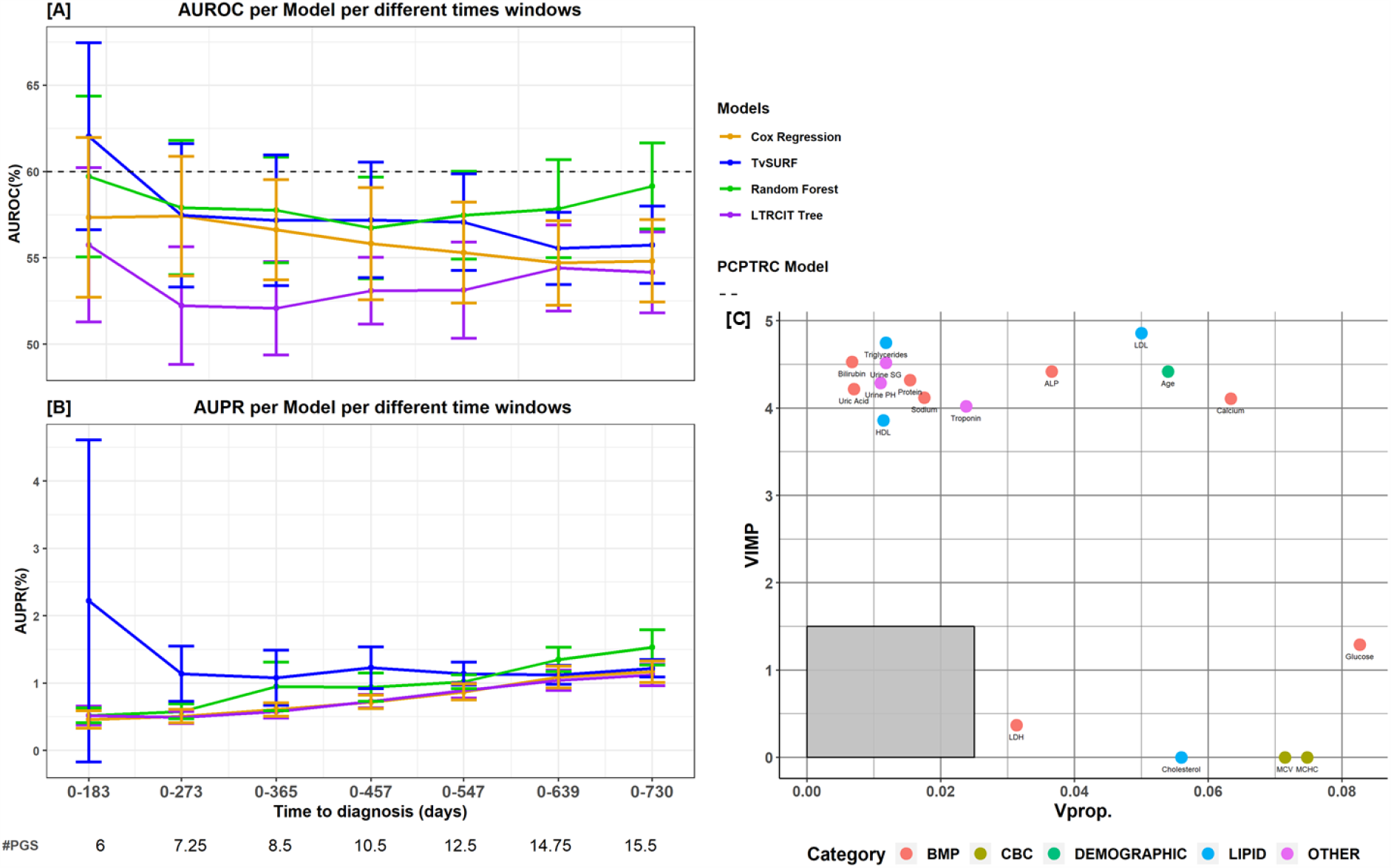
PGC risk prediction and variable importance. [A] Performance (AUROC mean±SD) of five prediction models for different time windows. The grey dashed line represents the (time-independent) AUROC previously reported for the PCPTRC model. [B] AUPR. The numbers below the x-axis labels are the average number of individuals with PGC that were available across the cross-validation folds for each time interval. [C] Variable importance for model prediction in a 183 day window. Points indicate the different variables. Axis definitions are as in Figure 6. The color of a point represents the variable’s category. Features of low importance (Vprop <0.025 and VIMP<1.5) are not shown.

#### Variable importance

**Figure 5C** summarizes the importance of the variables used by TVsuRF in PGC risk prediction, for the 183-day window. The covariates alkaline phosphatase (ALP), low-density lipoprotein (LDL), age, calcium, and glucose had the largest impact on the model. Most of the lipids that were measured - LDL, high-density lipoprotein (HDL), cholesterol and triglycerides - had high importance risk according to at least one criterion, in agreement with previous reports [48].

## DISSCUSSION

In this article, we introduced a method for survival prediction based on time-varying covariates utilizing an ensemble of survival trees, and applied it for predicting future emergence of breast and prostate cancer. Our method outperformed traditional prediction methods in breast cancer and for short term prediction also in prostate cancer. While traditional survival analysis methods use prior assumptions concerning the distribution of the data [49], our method relies only on the proportional-hazard assumption.

Our work has several limitations. First, we do not directly address the issue of size imbalance between the negative (here, the majority) and positive classes. That could affect the splitting criteria and produce nodes with a small number of samples or nodes without failure events, especially in datasets with high-dimensional feature space. Methods such as synthetic minority sampling might address this point [50]. Second, since our dataset did not record the existing clinical models for cancer risk (Gail’s model for BC, and PCRTRC model for PGC), we could not compare performance to them on individual patients in our cohort. Incorporating them as additional features in our models may improve prediction. Third, the small number of visits per patient did not allow us to incorporate into the model time-related features, as suggested, e.g., in [19,51] engineered features that capture interactions [52], or to model per-patient random effects across pseudo-intervals. Other model extensions such as competing risks (e.g. death) and accounting for cardiovascular background were not possible for lack of data. Moreover, the limited cohort size made it difficult to evaluate the calibration of our model.

Future work should examine different imputation methods, as those might affect the performance of classifiers when modeling EMR data [53], and investigate sequential models that incorporate the full history in predicting the personalized survival curve [54]. In addition, ‘out-of-bag’ approaches may improve the evaluation of the prediction, as previously suggested [55]. Moreover, the robustness of the approach is yet to be demonstrated on EMR data from other medical centers. Predictions for additional types of cancers should also be tested, given sufficient data. Finally, a prospective clinical study would provide a more accurate evaluation of the performance.

## CONCLUSIONS

Our models demonstrate the potential of using common laboratory tests of healthy individuals to assess cancer risk. They can serve as additional screening tests and complement the existing BC screening methods.

## Data Availability

Data cannot be shared for ethical/privacy reasons.

## ACKNOWLEDGEMENT

None.

## COMPETING INTERESTS

None.

## FUNDING

Supported in part by Israel Science Foundation (ISF) grant No. 1339/18 (RS); ISF grant No. 3165/19, within the Israel Precision Medicine Partnership program (RS); grant 2016694 from the US - Israel Binational Science Foundation (BSF), and the US National Science Foundation (NSF) (RS); ELROV grant (S.S.T). D.C. was supported, in part, by fellowships from the Edmond J. Safra Center for Bioinformatics at Tel Aviv University and from Google.

## DATA AVAILABILITY

Data cannot be shared for ethical/privacy reasons.

## Notes

### Competing Interest Statement

The authors have declared no competing interest.

### Author Declarations

Tel-Aviv Sourasky Medical Center Institutional Review Board (Approval no. 02-049-Tlv) - ethical approval was given.

