## Supplementary for "Early detection of prostate gland and breast cancer risk based on routine check-up data using survival analysis trees for left-truncated and right-censored data"

### Supplementary Material

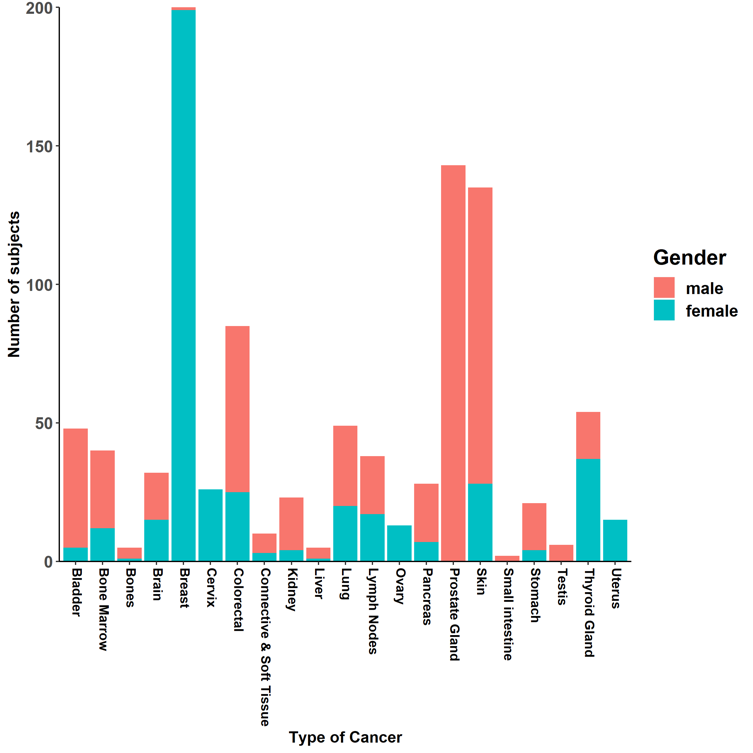

**Supplementary Figure 1: Number of patients per cancer type**.

Bar plot of the number of individuals who were surveyed in TAMICS and later diagnosed with cancer, categorized by gender and type of cancer.

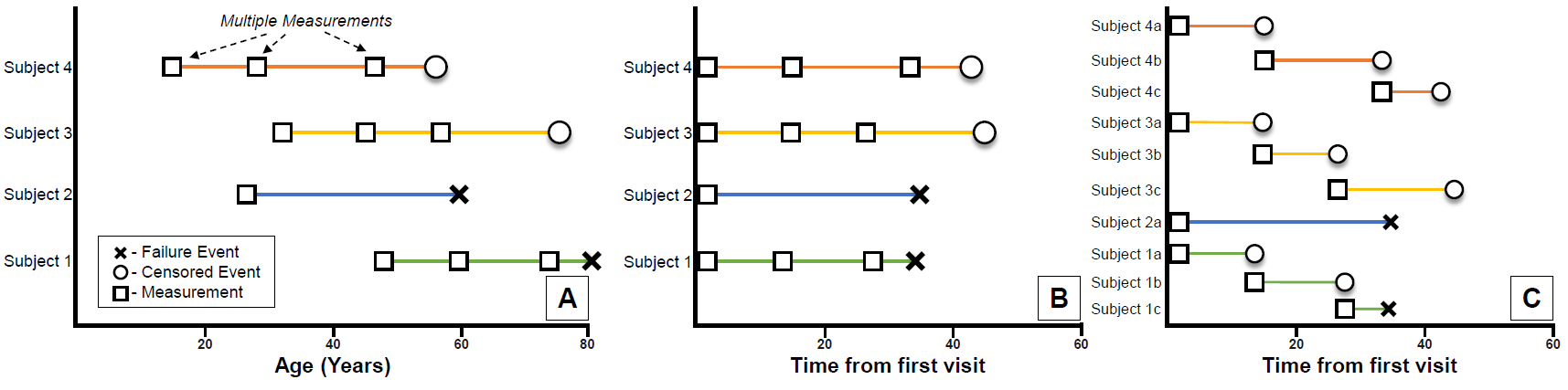

**Supplementary Figure 2:** **Presenting longitudinal data of multiple visits.**
(A) Longitudinal measurements and survival analysis setting. Squares indicate the times of the longitudinal measurements, Crosses indicate failure events, and circles indicate censoring events. (B) The data after shifting all first visit times to 0. (C) The same data after transforming into pseudo-objects.

**Supplementary Material 3: Decision tree basics**

Binary trees:

A rooted tree $T$ is a connected acyclic graph with a designated node $r$ called the *root*. Other nodes of degree $1$ are called *leaves*. In such a tree there is a single simple path from $r$ to every node and the number of edges in the path is the *depth* of the node. If there exists a simple path from $r$ to $v$ that passes through $u$then $u$is called an *ancestor* of $v$. If also $(u,v)$ is an edge then $u$ is the *father* of $v$ and $v$ is *child* of $u$. If every non-leaf has two children then $T$ is called a *binary tree*.

Decision trees:

A binary rooted tree can be used as a decision tree for classification as follows: Each internal (non-leaf) node is associated with a certain covariate and a threshold value. Samples with the covariate value above the threshold are assigned to the right child, and the rest are assigned to the left. This way, a sample starts at the root and descends left or right depending on the corresponding covariate values until it is associated with a leaf. If leaves are assigned with a class label (e.g. case / control), the tree assigns a class for the sample. Similarly, a set of samples can be partitioned into disjoint subsets corresponding to the leaves. Note that in our case the samples are the LTRC pseudo-intervals.

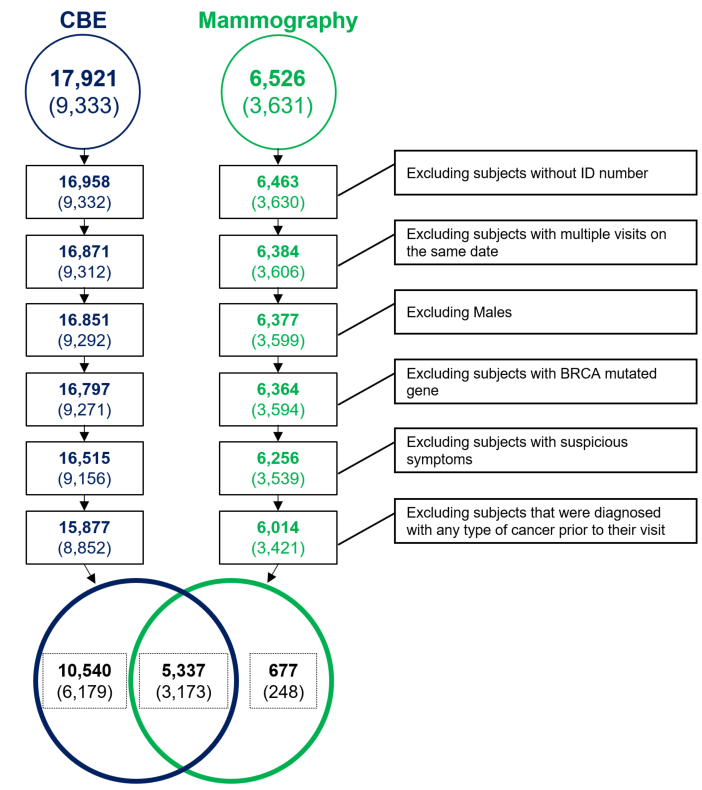

**Supplementary Figure 4**: **The CBE and mammography cohorts.** Effect of exclusion criteria on the members of the TAMICS cohort who conducted a mammography screening test for BC and CBE.

**Supplementary Material 5:** Cohort of subjects with CBE and Mammography tests.

We removed all the visits that occurred less than 31 days after the previous one. We excluded all subjects with two or more types of cancer unless the only other type was skin cancer. In case of more than one BC diagnosis we considered only the first one.

We used natural language processing to classify each subject who was recommended to conduct any BC-related follow-up test as positive (abnormal mammography). The extraction of the recommendation from the physician's notes was done using a pattern detection script. All phrases after an action verb, such as ‘is required’; ‘recommend’; were extracted and a dictionary of words that indicate BC follow-up test (ultrasound, biopsy, trucut etc.) was created. We manually reviewed the mammography results and added more action verbs and recommendations in several iterations. Finally, we randomly sampled and manually reviewed 100 cases to confirm the efficacy of our pattern recognition script.

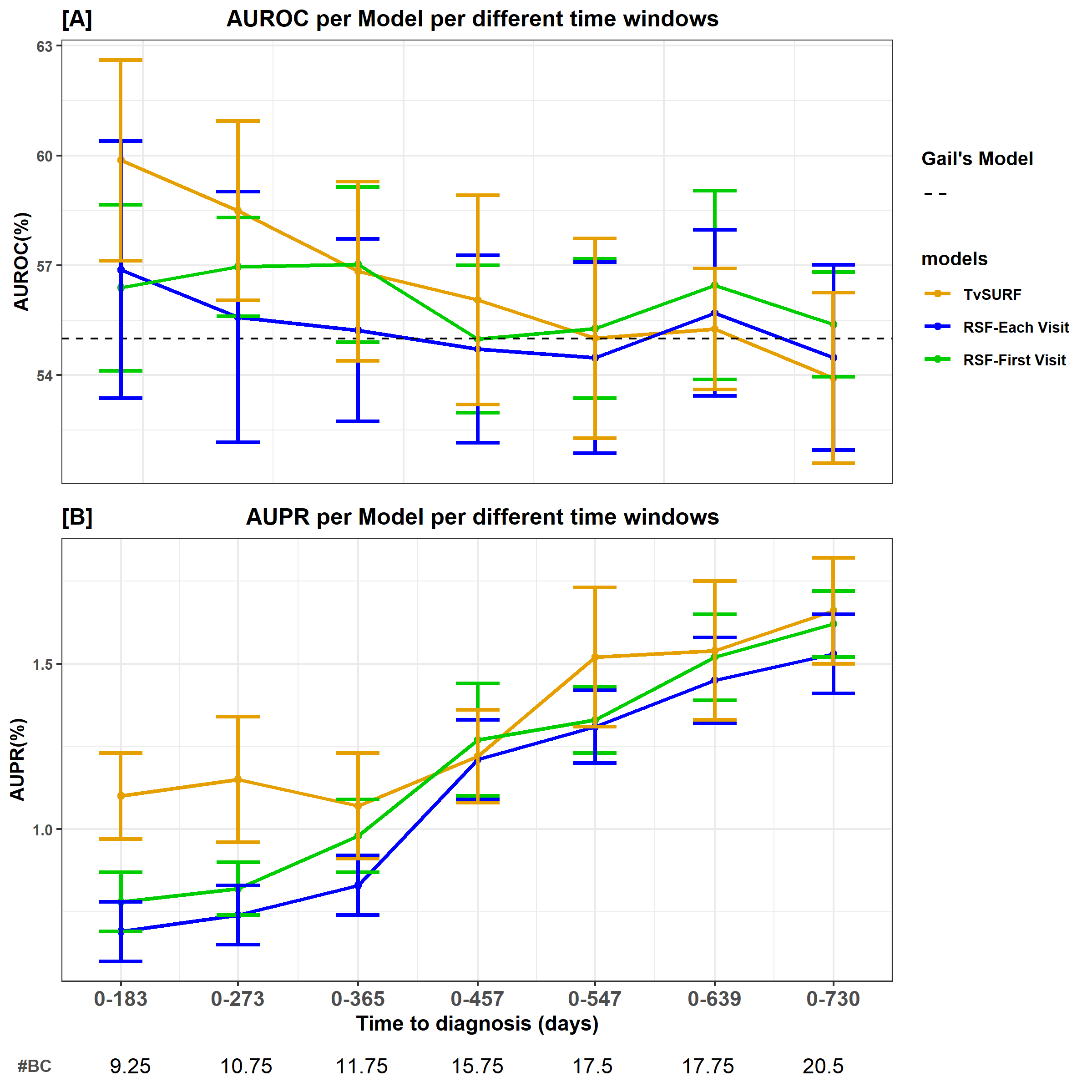

**Supplementary Figure 6:** **BC risk prediction – comparison of TVsuRF to random survival forest**. Two versions of RSF were applied: Each Visit: All pseudo-intervals were used. First Visit: Every visit creates an interval starting at the visit time and ending at the time of failure or censoring of the subject. In the two versions, all pseudo-intervals were linearly shifted to start at time t=0 since the RSF models are time independent. The numbers below the x-axis labels are the average number of BC patients that were available across the cross-validation folds for each time interval.

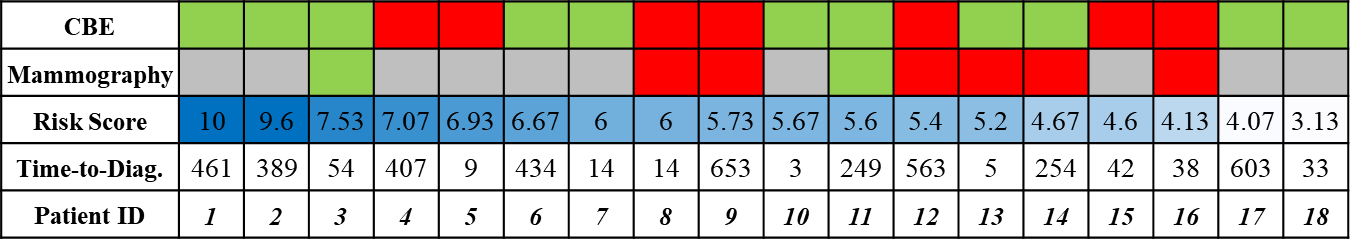

**Supplementary Figure 7**: **TVsuRF risk score and BC screening tests results for women who subsequently were diagnosed with BC**. Green: a normal result; Red: an abnormal test; Grey: test unavailable. 1^s^ line: CBE result, 2^nd^ line: mammography result; 3^rd^ line: the risk score calculated by the TVsuRF model. Patients were ordered from high (dark blue) to low (light blue) risk score. 4^th^ line: time from visit to cancer diagnosis.

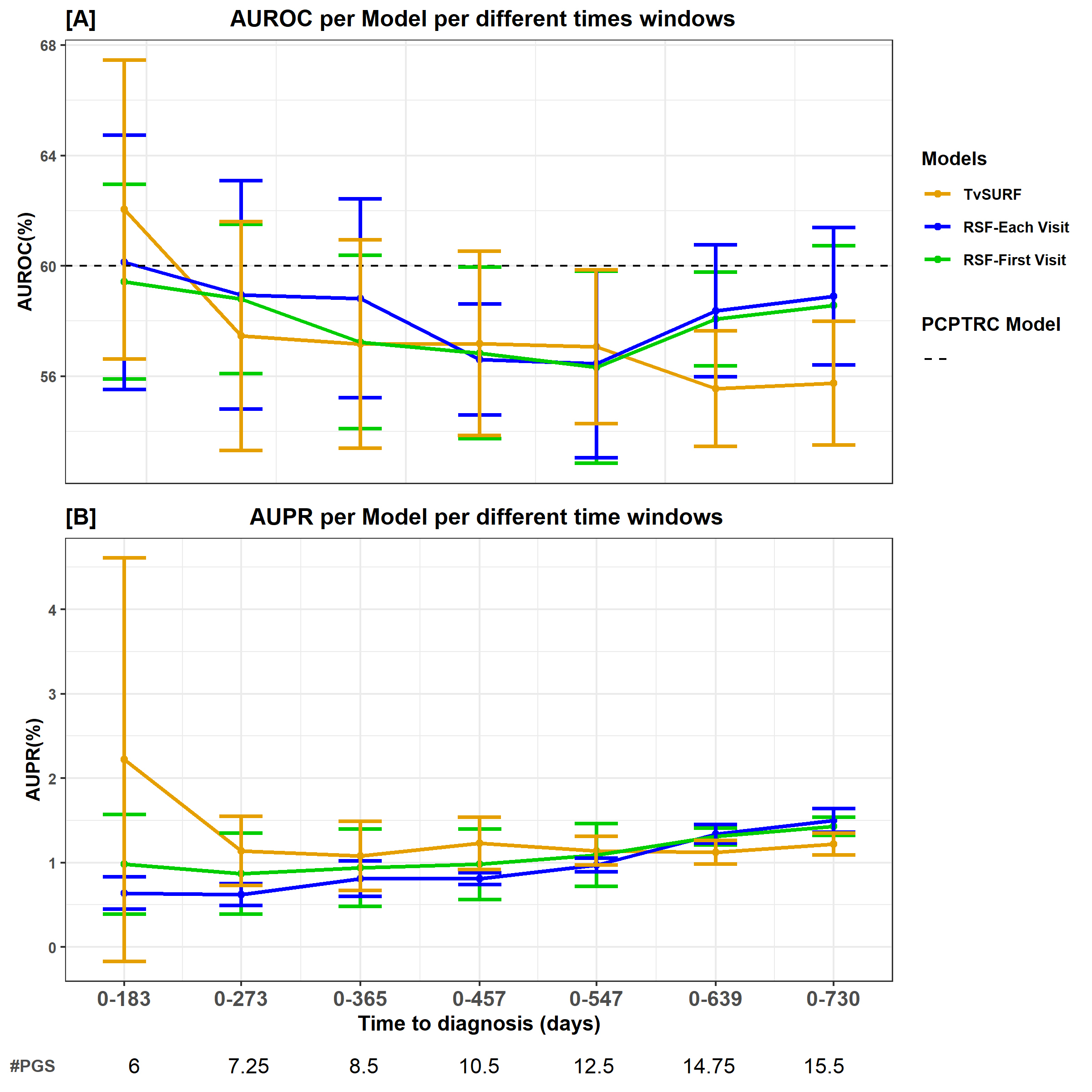

**Supplementary Figure 8:** **PGC risk prediction – comparison of TVsuRF to random survival forest.** The same two RSF variants in SFig. 6 were used. The grey dashed line represents the (time-independent) AUROC previously reported for the PCPTRC model.
